# A Fractal Viewpoint to COVID-19 Infection

**DOI:** 10.1101/2020.06.03.20120576

**Authors:** Oscar Sotolongo-Costa, José Weberszpil, Oscar Sotolongo-Grau

## Abstract

One of the central tools to control the COVID-19 pandemics is the knowledge of its spreading dynamics. Here we develop a fractal model capable of describe this dynamics, in term of daily new cases, and provide quantitative criteria for some predictions. We propose a fractal dynamical model using conformed derivative and fractal time scale. A Burr-XII shaped solution of the fractal-like equation is obtained. The model is tested using data from several countries, showing that a single function is able to describe very different shapes of the outbreak. The diverse behavior of the outbreak on those countries is presented and discussed. Moreover, a criterion to determine the existence of the pandemic peak and a expression to find the time to reach herd immunity are also obtained.

## 1. INTRODUCTION

The worldwide pandemic provoked by the SARS-CoV-2 coronavirus outbreak have attracted the attention of the scientific community due to, among other features, its fast spread. Its strong contamination capacity has created a fast growing population of people enduring COVID-19, its related disease, and a non small peak of mortality. The temporal evolution of contagion over different countries and worldwide brings up a common dynamic characteristic, in particular, its fast rise to reach a maximum followed by a slow decrease (incidentally, very similar to other epidemic processes) suggesting some kind of relaxation process, which we try to deal with, since relaxation is, essentially, a process where the parameters characterizing a system are altered, followed by a tendency to equilibrium values. In Physics, clear examples are, among others, dielectric or mechanical relaxation. In other fields (psychology, economy, etc.) there are also phenomena in which an analogy with “common” relaxation can be established. In relaxation, temporal behavior of parameters is of medular methodological interest. That is why pandemics can be conceived as one in which this behavior is also present. For this reason, we are interested, despite the existence of statistical or dynamical systems method, in the introduction of a phenomenological equation containing parameters that reflect the system’s behavior, from which its dynamics emerges. We are interested in studying the daily presented new cases, not the current cases by day. This must be noted to avoid confusion in the interpretation, i.e. we study not the cumulative number of infected patients reported in databases, but its derivative. This relaxation process in this case is, for us, an scenario that, by analogy, will serve to model the dynamics of the pandemics. This is not an ordinary process. Due to the concurrence of many factors that make very complex its study, its description must turn out to non classical description. So, we will consider that the dynamics of this pandemic is described by a “fractal” or internal time [1]. The network formed by the people in its daily activity forms a complex field of links very difficult, if not impossible, to describe. However, we can take a simplified model where all the nodes belong to a small world network, but the time of transmission from one node to other differs for each link. So, in order to study this process let us assume that spread occurs in “fractal time” or internal time [1, 2]. This is not a new tool in physics. In refs. [3-5] this concept has been successfully introduced and here, we keep in mind the possibility of a fractal-like kinetics [6], but generalizing as a nonlinear kinetic process. Here we will follow to what we refer as a “relaxation-like” approach, to model the dynamics of the pandemic and that justify the fractal time. By analogy with relaxation, an anomalous relaxation, we build up a simple nonlinear equation with fractal-time. We also regain the analytical results using a deformed derivative approach, using conformable derivative (CD) [7]. In Ref. [8] one of the authors (J.W.) have shown intimate relation of this derivative with complex systems and nonadditive statistical mechanics. This was done without resort to details of any kind of specific entropy definition.

Our article is outlined as follows: In Section 2, we present the fractal model formulated in terms of conformable derivatives, to develop the relevant expressions to adjust data of COVID-19. In Section 3. we show the results and figures referring to the data fitting along with discussions. In section 4, we finally cast our general conclusions and possible paths for further investigations.

## 2. FRACTAL MODEL

Let us denote by *F(t)* the number of contagions up to time *t*.

The CD is defined as [7]

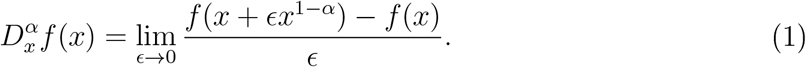

Note that the deformation is placed in the independent variable.

For differentiable functions, the CD can be written as

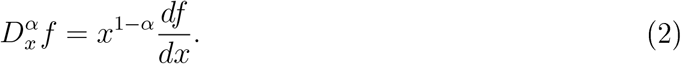

An important point to be noticed here is that the deformations affect different functional spaces, depending on the problem under consideration. For the conformable derivative [8-12], the deformations are put in the independent variable, which can be a space coordinate, in the case of, e.g, mass position dependent problems, or even time or spacetime variables, for temporal dependent parameter or relativistic problems. Since we are dealing with a complex system, a search for a mathematical approach that could take into account some fraetalitv or hidden variables seems to be adequate. This idea is also based in the fact that we do not have full information about the system under study. In this case, deformed derivatives with fractal time seems to be a good option to deal with this kind of system. Deformed derivatives, in the context of generalized statistical mechanics are present and connected [8]. There, the authors have also shown that the *q – deformed* derivative has also a dual derivative and a *q – exponential* related function [13]. Here, in the case under study, the deformation is considered for the solutions-space or dependent variable, that is, the number *F(t)* of contagions up to time *t*. One should also consider that justification for the use of deformed derivatives finds its physical basis on the mapping into the fractal continuum [8, 14-16]. That is, one considers a mapping from a fractal coarse grained (fractal porous) space, which is essentially discontinuous in the embedding Euclidean space, to a continuous one [9]. In our case the fractality lies in the temporal variable. Then the CD is with respect to time.

A nonlinear relaxation model can be proposed here, again based on a generalization of Brouers-Sotolongo fractal kinetic model (BSf) [3, 4, 17], but here represented by a nonlinear equation written in terms of CD:

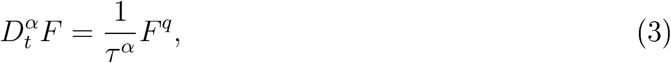

where *τ* is our “relaxation time” and *q* and *α* here are real parameters. We do not impose any limit for the parameters. Equation (3) has as a well known solution a function with the shape of Burr XII [18], with:

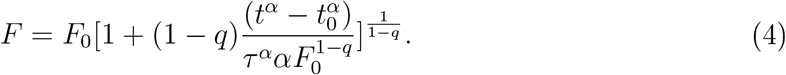

The density (in a similar form of a PDF, but here it is not a PDF) is, then:

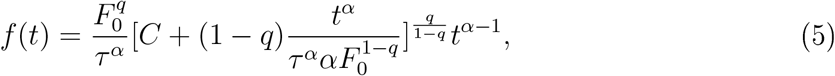

where 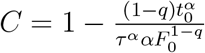, which can be expressed as:

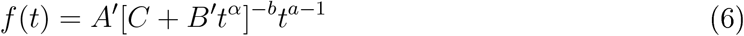

where the parameter are 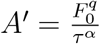*, 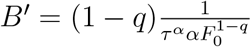*, 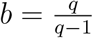, *a = α*.

Or, in a simpler form for data adjustment purposes

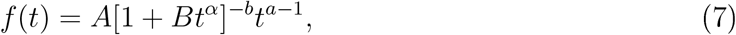

with 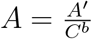*, 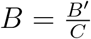*.

This is very similar, though not equal, to the function proposed by Tsallis [19, 20] in an ad hoc way. Here, however, a physical representation by the method of analogy is proposed to describe the evolution of the pandemics. Though we have introduced *A, B, C*, b, and *a* as parameters to simplify the fitting, the true adjustment constants are, clearly, *q, τ* and *α*. Note that we do not impose any restrictive values to the parameters.

There is no need to demand that the solution always converge. The equation to obtain Burr XII has to impose restrictions but this is not the case. In Burr XII the function was used as a probability distribution. But here the function describes a dynamic, which can be explosive, as will be shown for the curves of Brazil and Mexico. Therefore, if we consider infinite population, a peak will never be reached unless the circumstances change (treatments, vaccines, isolation, etc.). Our model does not impose finiteness of the solution. The possibility for a decay of the pandemic in a given region in this model requires the fulfillment of the condition

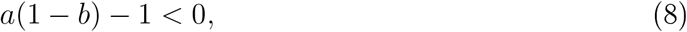

what expresses the property that

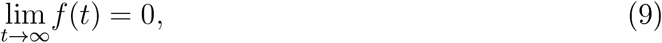

what means that the function has a local maximum. If this condition is not accomplished, the pandemic does not have a peak and, therefore, the number of cases increases forever in this model.

In this case there is, apart from the change of propagation and development conditions, the possibility for a given country that does not satisfies condition (8), to reach “herd immunity”, i.e., when the number of contagions has reached about 60% of population, in

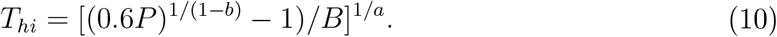

We will work with what we will call *T*_1000_ ahead and that seems to make more sense and bring more information.

## 3. DATA FITTING

With eq. (7) let us fit the data of the epidemic worldwide. The data was extracted from Johns Hopkins University [21] and the website [22] to process the data for several countries.

We covered the infected cases taken at Jan 22 as day 1, up to June 13. The behavior of new infected cases by day is shown in figure 1. The fitting was made with gnuplot 5.2. As it seems, the pandemic shows some sort of “plateau”, so the present measures of prevention are not able to eliminate the infection propagation in a short term, but it can be seen that condition (8) is weakly fulfilled.

**Figure 1.**
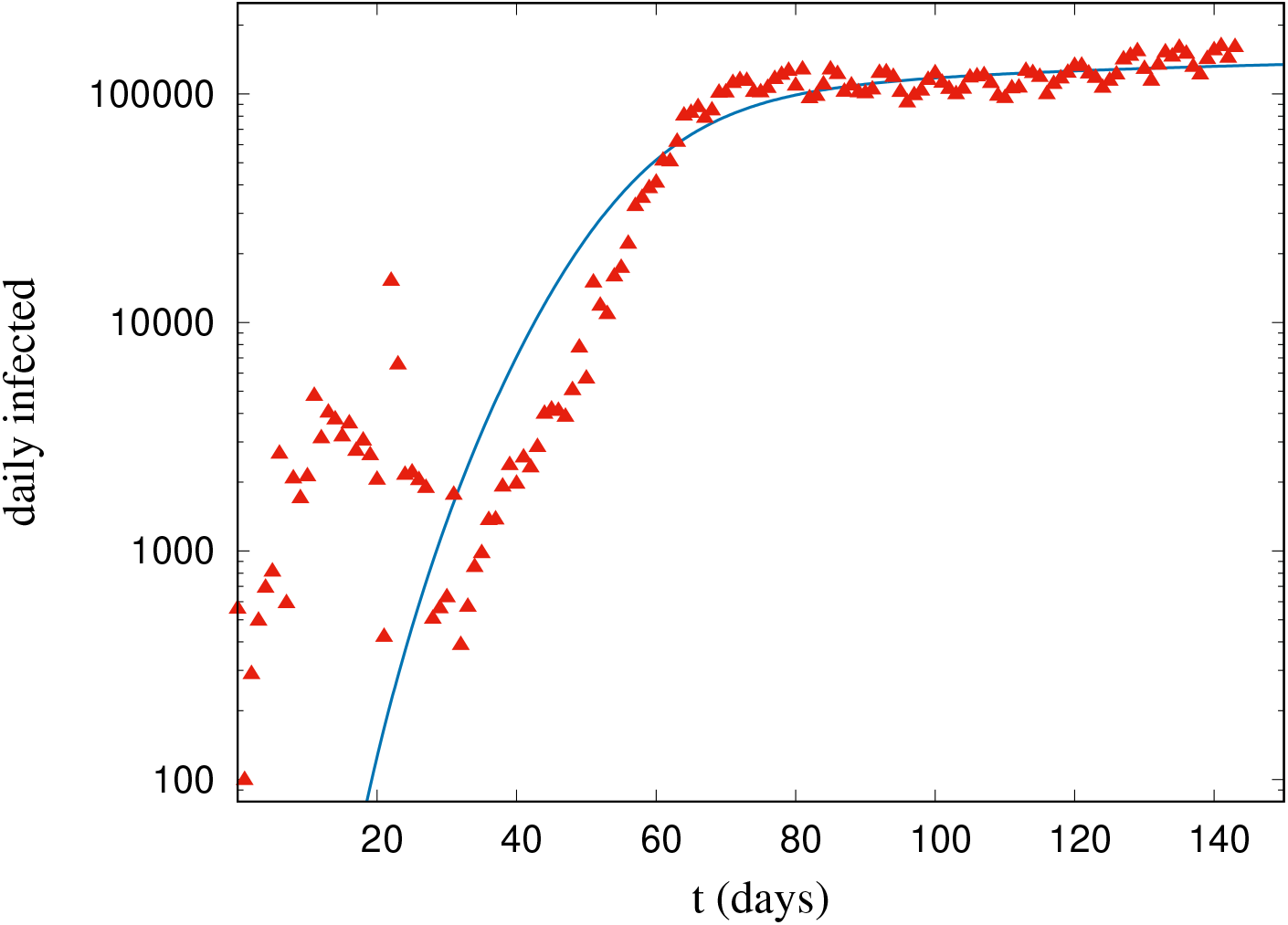
Worldwide infections from Jan, 22 to June 13 and fitting with eq. (7). The behavior fits well with parameters in Table I. Condition (8) is satisfied.

In the particular case of Mexico the fitting is shown in figure 2. In this case condition (8) is not fulfilled. In terms of our model this means that the peak is not predictable within the present dynamics. Something similar occurs with Brazil, as shown in figure 3. The data for Brazil neither fulfill the condition (8). In this case there is neither the prevision of a peak and we can say that the data for Mexico and Brazil reveals a dynamics where the peak seems to be quite far if it exists. But there are some illustrative cases where the peak is reached. Progression of the outbreak in Cuba and Iceland are shown in Figure 4 and 5 respectively. Condition (8) is satisfied for both countries and we can see that the curve of infection rate descends at a good speed after past the peak. How let us take a look at United States data, shown in Figure 6. The USA outbreak is characterized by a very fast growth until the peak and, then, very slow decay of the infection rate is evident. As discussed above, the outbreak will be controlled for almost infinite time in this dynamics. There is also some intermediate cases as Spain and Italy, shown in Figures 7 and 8. In this case the data exhibits the same behavior as in USA, a fast initial growth and a very slow decay after the peak. However, the outbreak is controlled in a finite amount of time. In Table I we present the relevant fitting parameters, including herd immunity time, *T_hi_* and *T*_1000_, the time to reach a rate of 1000 infections daily. This, for countries that have not reached the epidemic peak, Mexico and Brazil, We also include the population, P; of each country.

**Figure 2.**
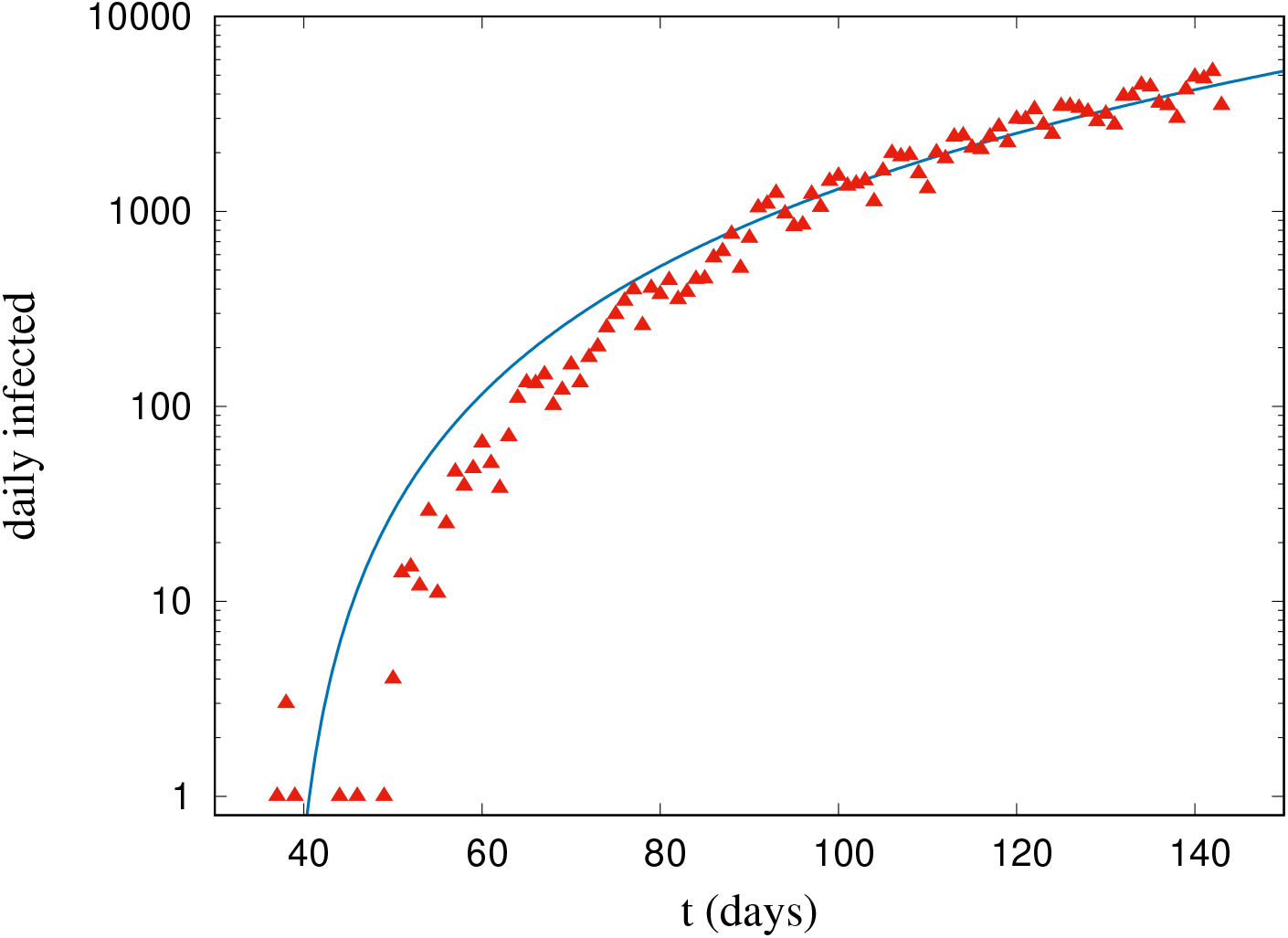
Daily infections in Mexico and fitting with eq. (7) for parameters in Table I. *T_hi_ =* 778 days. Condition (8) is not satisfied.

**Figure 3.**
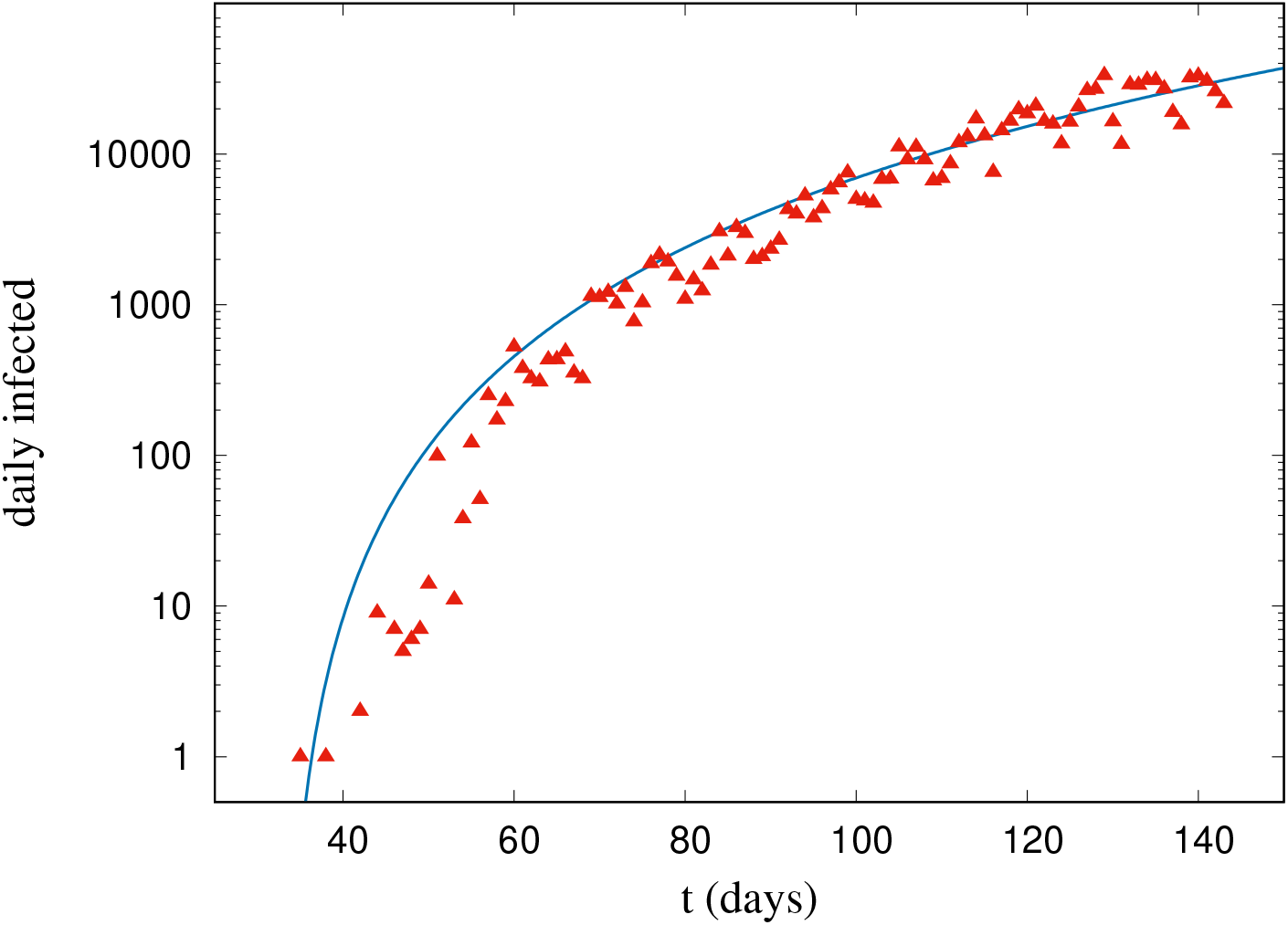
Evolution of daily cases in Brazil and fitting with eq. (7) for parameters in Table I. *T_hi_ =* 298 days. Condition (8) is not satisfied.

**Figure 4.**
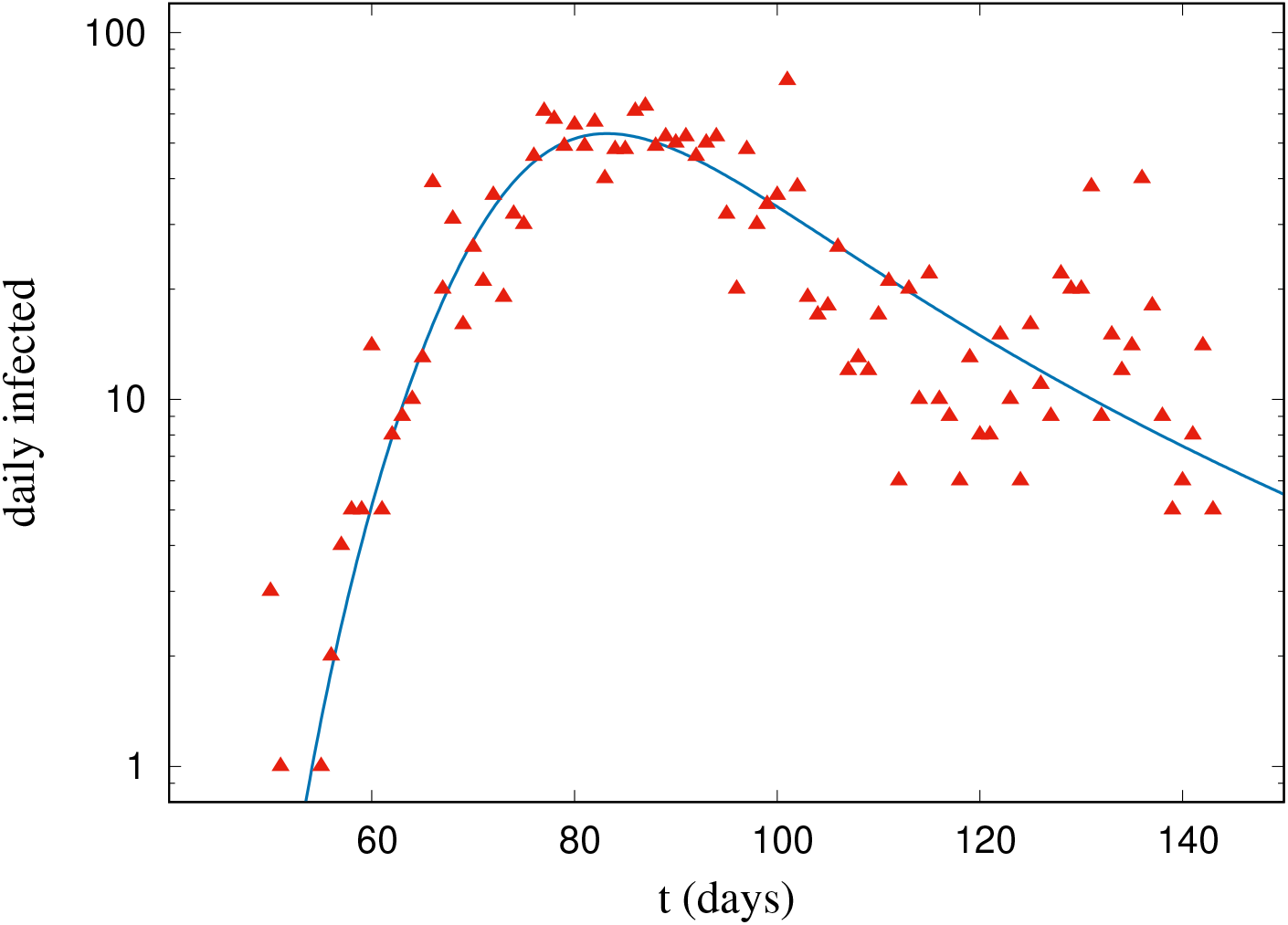
Daily infections in Cuba. The theoretical curve fits with data though with a poor correlation due to the dispersion. See fitting parameters in Table I. Condition(8) is satisfied.

**Figure 5.**
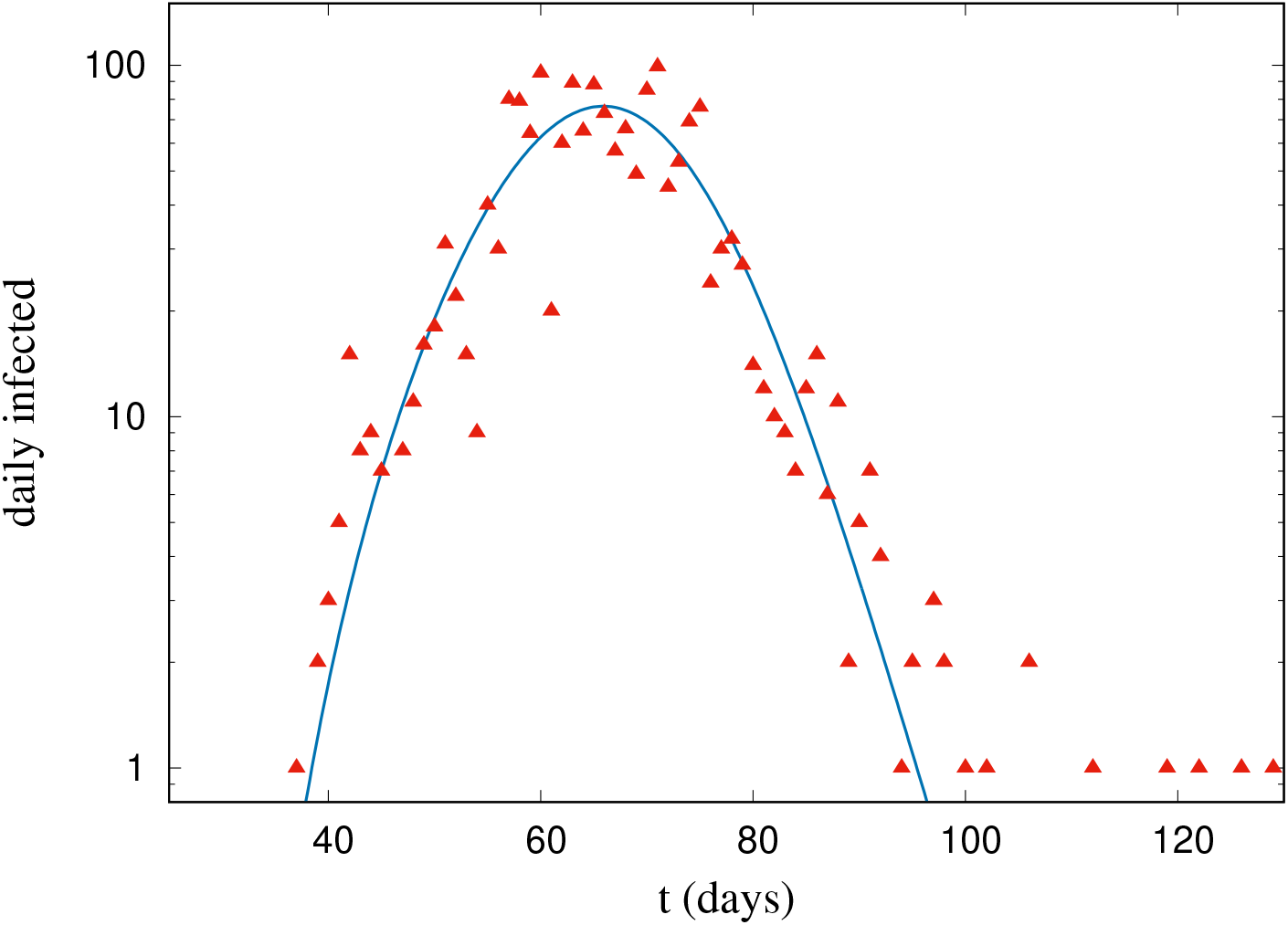
Daily infections in Iceland, where the pandemic seems to have ceased. Here again, in spite of the relatively small correlation coefficient, the behavior of the pandemic in this country looks well described by eq. (7). See fitting parameters in Table I. Condition (8) is satisfied.

**Figure 6.**
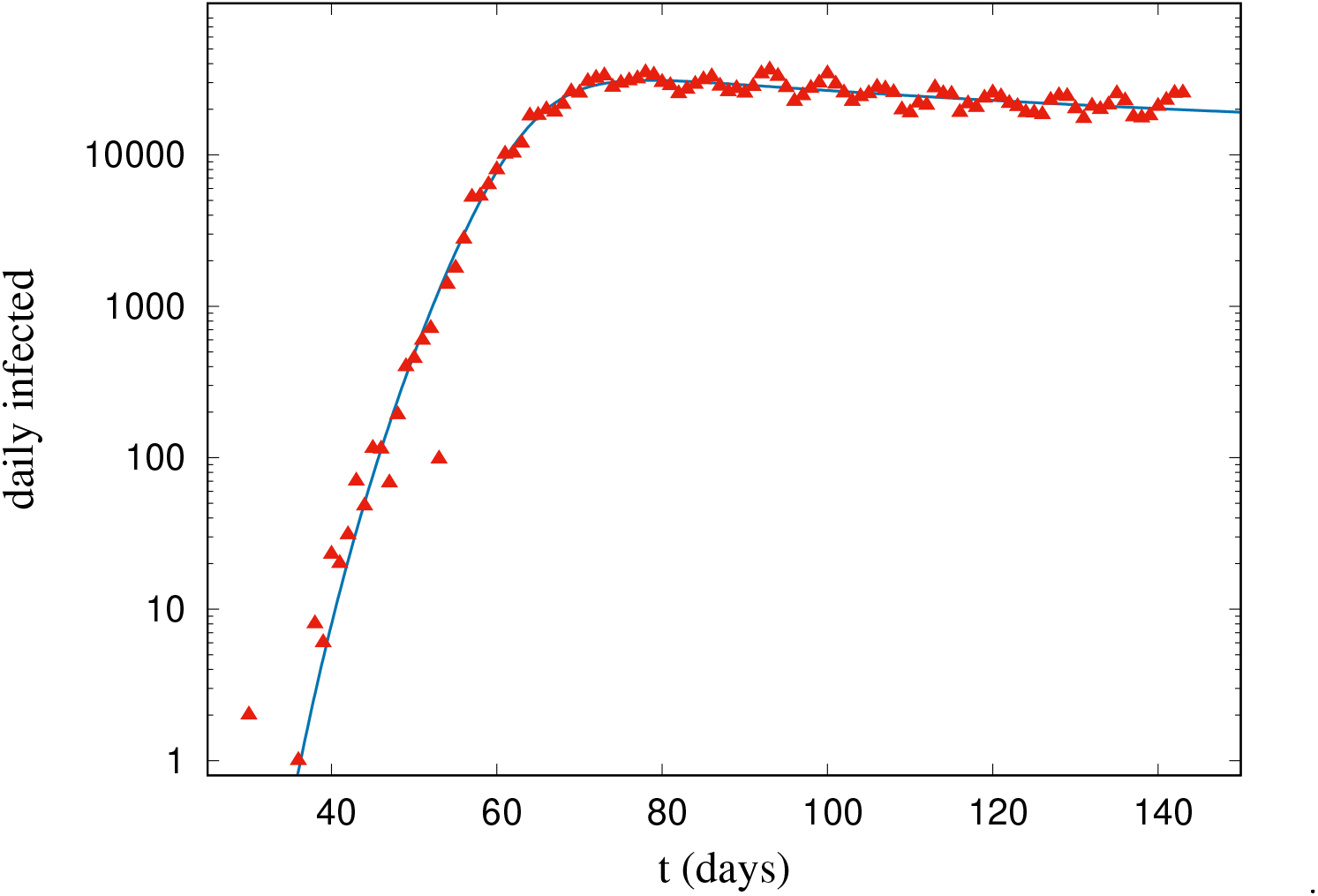
Daily infections in USA, where the peak looks already surpassed. Here again, the behavior of the pandemic in this country looks well described by eq. (7). See fitting parameters in Table I. Condition (8) is satisfied.

**Figure 7.**
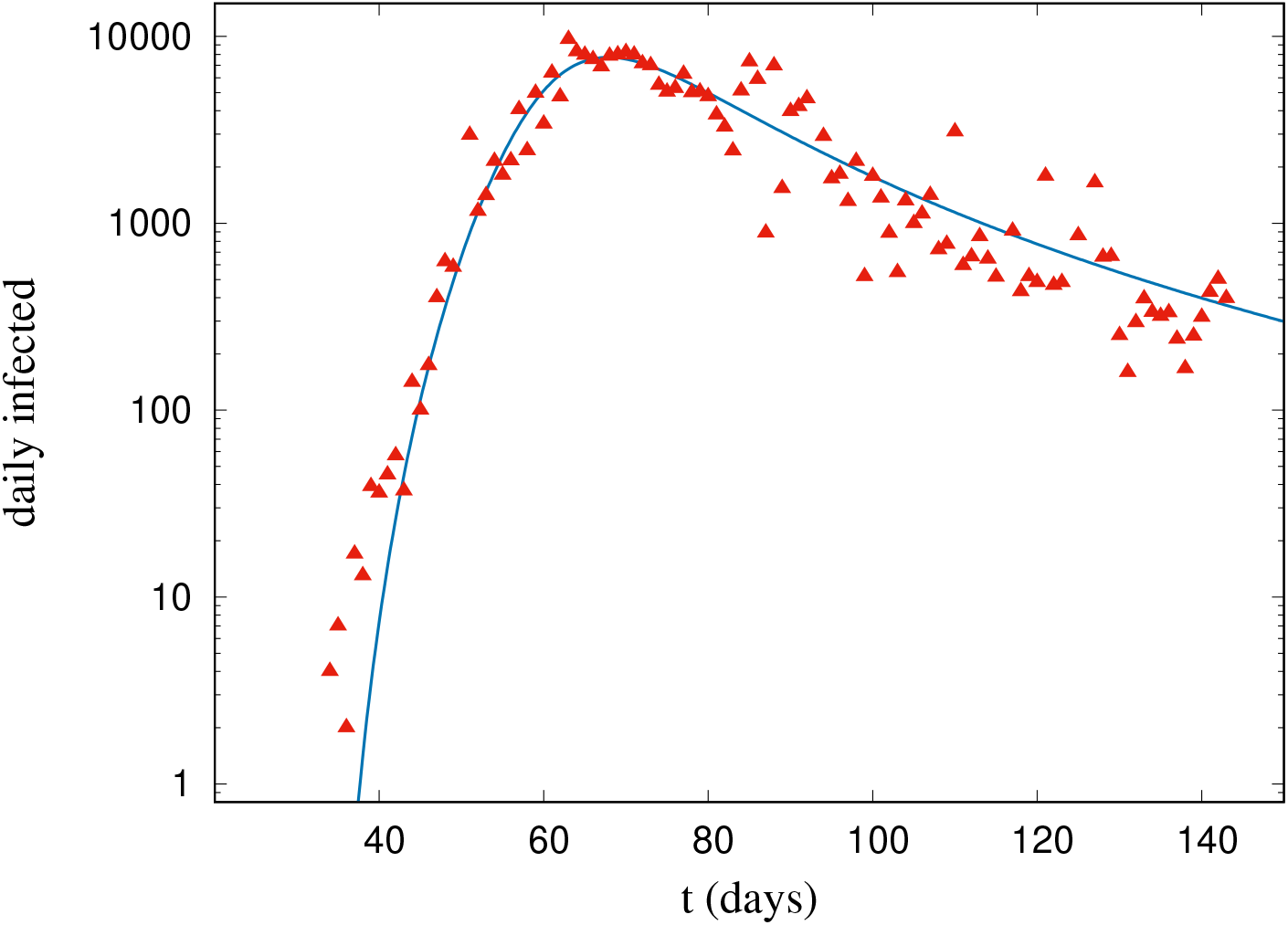
Daily infections in Spain. the data shows a large dispersion but the curve describes well the behavior. See fitting parameters in Table I. Condition (8) is satisfied.

**Figure 8.**
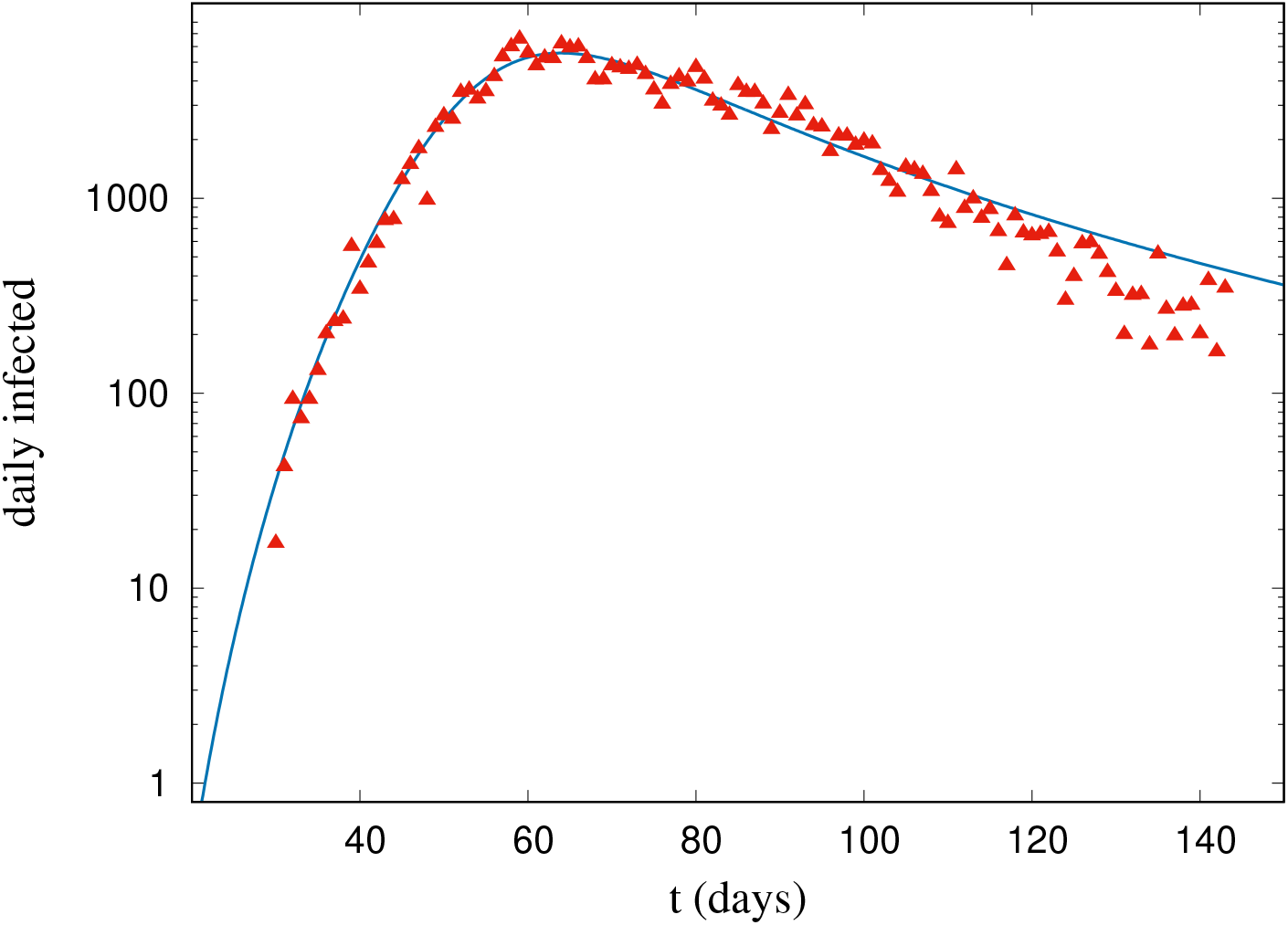
Daily infections in Italy. See fitting parameters in Table I. Condition (8) is satisfied.

**Table I.** Relevant fitting parameters for each country.

| Country | A | B | a | b | P | $T_{hi}$(days) | $T_{1000}$ |
| --- | --- | --- | --- | --- | --- | --- | --- |
| Brazil | 0.0152828 | 0.0104434 | 4.31197 | 0.0671095 | 212559417 | 298 | 36 |
| Cuba | 1.80E-05 | 3.30E-09 | 5.31906 | 1.40779 | 11326616 | - | - |
| Iceland | 6.08E-05 | 1.69E-09 | 5.09845 | 4.94326 | 341243 | - | - |
| Italy | 1.20E-07 | 1.85E-13 | 7.50956 | 1.32858 | 60461826 | - | 34 |
| Mexico | 0.0541958 | 0.0104956 | 3.60005 | 0.0641971 | 128932753 | 778 | 56 |
| Spain | 0.000170317 | 2.75E-10 | 6.31706 | 1.35476 | 46754778 | - | 19 |
| USA | 1.09E-13 | 5.99E-20 | 11.5099 | 0.973087 | 331002651 | - | 34 |
| Worldwide | 3.18E-06 | 4.65E-13 | 6.84834 | 0.816744 | 7786246434 | - | 29 |

As can be seen from fitting coefficients, the exponent *b* drives the behavior of infections in every country. Those countries that manage well the disease expansion have b values wide larger than 1. Countries with b values close to one, as Italy and Spain, have managed the pandemics but poorly and at high costs. The recovery in both countries will be long. The same is valid for USA, that manage poorly the outbreak and its struggling with an even longer recovery to normal life. Even worst scenario is taken place in Mexico and Brazil, with very low values of *b*. Those countries are experiencing a big outbreak where even can get herd immunity This, however, implies very high values of infections and mortality for the near future.

But let us briefly comment about herd immunity. Those countries that have managed to stop the outbreak, even with relative high mortality as Spain and Italy, will not reach the herd immunity As a matter of fact. This can not be calculated for those countries. Then, we can see countries like Brazil where, if the way of deal with the outbreak do not change, the herd immunity will be reached. Even when it seems desirable, the ability to reach the herd immunity brings with it a high payload. That is, for a country like Brazil the herd immunity would charge more than 100 million of infected people. That is, much the same as if a non small war devastates the country There is an alike scenario in Mexico, but the difference here is that the value for *T_hi_* is so high that SARS-CoV-2 could even turn into a seasonal virus, at least for some years. We can expect around the same mortality but scattered over a few years.

A special observation deserves USA, where *T_hi_* tends to infinity. Here we can expect a continuous infection rate for a very long time. The outbreak is controlled but not enough to eradicate the virus. Virus will not disappear in several years but maybe the healthcare system could manage it. The virus will get endemic, and immunity will never be reached. However the infections and mortality rate associated with it, can be, hypothetically, small if compared with Mexico and Brazil. We can also compare the speed of the outbreak in different countries. As we already said in Table I we calculated *T*_1000_ for some countries. However, it should be noticed that this time is not calculated from day 0, which is always January 22, but for the approximated day when the outbreak began in the correspondent country. By example, in Brazil there was no cases at January, 22 but the first cases were detected around March, 10. So both, data fitting and *T*_1000_, were calculated from March, 10.

## 4. CONCLUSIONS AND OUTLOOK FOR FURTHER INVESTIGATIONS

In this work, for the first time, we presented a model built using the method of analogy, in this case with a nonlinear relaxation-like behavior. With this, a good fitting with the observed behavior of the daily number of cases with time is obtained. The explicit expressions obtained may be used as a tool to approximately forecast the development of the COVID-19 pandemic in different countries and worldwide. In principle, this model can be used as a help to elaborate or change actions. This model does not incorporate any particular property of this pandemic, so we think it could be used to study pandemics with different sources. With the collected data of the pandemics at early times, using this model, it can be predicted the possibility of a peak, indefinite growth, time for herd immunity, etc.

What seems to be clear from the COVID-19 data, the fitting and the values shown in the Table I, is that SARS-CoV-2 is far from being controlled at world level. Even when some countries appear to control the outbreak, the virus is still a menace for its health system. Furthermore, in the nowadays interconnected world it is impossible for any country to keep closed borders and pay attention to what happens only inside. All isolation measures should be halted at some time and we can expect new outbreaks in countries like Spain or Italy even after the current one could be controlled. The only way to control the spread of SARS-CoV-2 seems to be the development of a vaccine that provides the so much desired herd immunity. Indeed, the model made possible to make an approximate forecast of the time to reach the herd immunity. This may be useful in the design of actions and policies about the pandemic. We have introduced the *T*_1000_, that gives information about the early infection behavior in populous countries. A possible improvement of this model is the formal inclusion of a formulation including the dual conformable derivative [12, 13]. This will be published elsewhere,

## Data Availability

All data from:
https://coronavirus.jhu.edu/data/new-cases.
https://www.worldometers.info/.to.

https://www.medrxiv.org/content/early/2020/05/04/2020.04.24.20078154.full.pdf.

https://coronavirus.jhu.edu/data/new-cases.

https://www.worldometers.info/.to.

## ACKNOWLEDGMENTS

We acknowledge Dr. Carlos Trallero -Giner for helpful comments and suggestions

